# CD206 upregulation in monocytes within whole blood cultures correlates with lung function in Cystic Fibrosis: a pilot study

**DOI:** 10.1101/2023.11.15.23298577

**Authors:** Sonali Singh, Jessica Longmate, David Onion, Paul Williams, Miguel Camara, Alan Smyth, Helen Barr, Luisa Martinez-Pomares

**Author notes:** Corresponding author: Luisa Martinez-Pomares.

## Abstract

Chronic inflammation dominates disease pathogenesis in Cystic Fibrosis (CF) and there is a need to characterise CF immunity. Whole blood cultures offer a cost-effective and non-invasive approach to investigate immune responses within the host environment. Here we used whole blood cultures to investigate the differentiation potential of monocytes (CD45+CD14+ cells) in CF (N=10) and controls (N=8) in the presence and absence of exogenous macrophage-colony stimulatory factor (M-CSF) or granulocyte-macrophage (GM)-CSF with and without interleukin (IL)-4. In CF and control cultures, CD45+CD14+ cells upregulated HLA-DR expression in all instances, and increased CD206 in the presence of GM-CSF with and without IL-4, and CD209 in the presence of GM-CSF and IL-4. In CF, we consistently observed reduced upregulation of CD206 in response to GM-CSF and a positive correlation between CD206 expression and lung function (FEV_1_). This was unique to cultured monocytes, and not seen with any other marker. These results highlight the potential of whole blood cultures to reveal cellular characteristics in differentiating monocytes related to clinical parameters that could guide the identification of novel biomarkers in CF.

## 1. Introduction

In Cystic Fibrosis (CF), chronic lung infection adversely affects life expectancy and quality of life. Pulmonary colonisation with pathogens such as *Pseudomonas aeruginosa*, negatively impacts lung function and can drive chronic neutrophilic inflammatory responses, leading to lung damage [1]. A tendency towards Th2/Th17-dominated immunity has described in CF [2]. There is a plethora of approaches to profiling human immunity with sample availability and processing being major factors to consider. Serum measurements can indicate clinical improvement in CF, for instance antibiotic treatment after exacerbation led to a reduction in circulating white blood cells and serum C-reactive protein, IL-6 and calprotectin [3]. Using stimulated peripheral blood mononuclear cells (PBMCs) we previously demonstrated that in people with CF IFN-γ production in response to phytohemagglutinin positively correlated with lung function (FEV_1_), whereas IL-17A production showed a negative correlation [4].

Monocytes are highly versatile and responsive immune cells that drive and respond to inflammatory processes and their characterisation could signpost inflammatory status in patients. Previous work identified changes in CF macrophages differentiated from purified blood monocytes compared to controls [5]. In addition, CF monocyte function is recovered in response to new therapeutic treatments [6, 7], which makes these cells possible therapeutic targets or prognosis markers in CF. Accordingly, monocytes driving neutrophilic inflammation in CF has recently been described [8].

In this study, we used whole blood cultures to identify potential differences in the phenotype of CF and control monocytes as they mature exposed to the host environment. This approach avoids potential artefacts caused by the cell purification process, reduces the sample size required, and takes into consideration the influence of plasma components and other blood cells. Whole blood cultures have been previously used to define immune status in volunteers [9, 10], and to establish how immune cells alter their phenotype under different culture conditions including treatment with SARS-CoV2 spike protein [11].

We analysed monocyte differentiation in the absence or presence of macrophage (M)-colony stimulating factor (CSF, CSF-1) or granulocyte-macrophage (GM)-CSF (CSF-2). Both factors contribute to macrophage homeostasis, with M-CSF produced constitutively and GM-CSF largely produced under inflammatory conditions [12]. GM-CSF also has an important homeostatic role in mucosal sites such as the lung where it is essential for the development of alveolar macrophages [13]. Although GM-CSF is widely considered to drive monocytes towards a pro-inflammatory phenotype[12], recent work analysing changes in gene expression and epigenetic modifications by Dabritz *et al*. indicate a potential regulatory function for GM-CSF-monocytes in the context of intestinal inflammation [14]. GM-CSF together with interleukin (IL)-4 was also included because of its ability to induce a dendritic cell-like phenotype in monocytes (CD206+CD209+) and confirm that IL-4 would be able to transduce a signal, if present [15, 16].

In this proof of principle study, we focused on cell activation markers (CD11b, HLA-DR) [17] and two lectin immune receptors (CD206 and CD209) shown to bind *P. aeruginosa* biofilms (CD206 and CD209) and planktonic cells (CD209) [16]. Our results show that monocytes upregulated HLA-DR, CD206, and CD209 upon culture and that CD206 expression in blood cultures positively correlates with FEV_1_, suggesting that monitoring selected surface markers in differentiating monocytes could inform on immune responses linked to lung pathophysiology in CF.

## 2. Materials and Methods

### Ethics

This study was approved by the Health Research Authority, REC reference: 17/LO/117, IRAS project ID: 190057. All participants’ details are presented in Table 1. None of the participants in the CF group were taking CFTR modulators.

**Table 1.** Demographic details for CF and matched control donors utilised in this study.

|  | **CF** | **Controls** |
| --- | --- | --- |
| **Number (n)** | 10 | 8 |
| **Sex (F/M)** | 4/6 | 3/5 |
| **Mean age ± SD (years)** | 32.4 ± 13.1 | 37.3 ± 13.5 |
| **Genotype** | ΔF508 homozygous: 7<br>ΔF508 heterozygous: 3 | N/A |
| **Median FEV<sub>1</sub> (range)** | 66.5 % (25%-109%) | N/A |
| **Positive *P. aeruginosa* culture** | Yes: 3<br>No: 7 (of which 5 have never been positive for PA) | N/A |
| **Azithromycin** | 4 | N/A |
| **Steroids (inhaled)** | 8 | N/A |
| **Steroids (oral)** | 1 | N/A |
| **Percentage of CD45+CD14+ cells in fresh blood (mean ± SEM)** | 6.94 ± 0.77 | 7.95 ± 0.74 |

### Reagents

Details of reagents used are given in Supplementary Table 1.

### Antibody staining of fresh blood

Whole blood was collected in citrate vacutainers. Samples (100 μl each) were incubated with relevant antibodies as per the manufacturer’s instructions for 15 min as follows: i. No antibodies (Unstained); ii. Isotype controls for CD45 and CD14; iii. Anti-CD45 and CD14 antibodies + isotype controls for HLA-DR, CD11b, CD206, and CD209; and iv. Anti-CD45, CD14, HLA-DR, CD11b, CD206, and CD209 antibodies (Supplementary Table 2). Samples were treated with OptiLyse C and processed as per the manufacturer’s instructions, resuspended in 0.1% paraformaldehyde in PBS, and stored at 4°C until analysis.

### Whole blood cultures

Blood was gently mixed and diluted 1:1 with warm RPMI complete medium (RPMI-1640 medium containing 15% human AB serum, 2 mM GlutaMAX, and 10 mM HEPES). The blood-medium mixture was cultured in 24-well ultra-low attachment plates (400 μl per well). Samples were either left untreated or treated with M-CSF (50 ng/ml), GM-CSF (20 ng/ml), or GM-SCF plus IL-4 (20 ng/ml each). For each donor, all four conditions were performed in duplicate. Cultures were incubated at 37°C, 5% CO_2_ for 24 h. Samples were labelled and processed for flow cytometry as for fresh blood.

### Flow analysis and data analysis

All samples were analysed on a MoFlo Astrios EQ flow cytometer within 24 h. The lymphocyte gate based on Forward and Size Scatter characteristics, was used to normalise for number of events (100,000 per fresh blood sample, 50,000 per 24 h cultured sample). Raw data were processed using Kaluza and further analysed using GraphPad Prism. Results are reported as the percentage of cells positive for each marker, as well as the relative median fluorescence intensity (rMFI) of the marker (rMFI=MFI of positive cells in stained sample/MFI of all cells in isotype control sample). Statistical tests are detailed in main text and figure legends, p<0.05 was considered significant.

## 3. Results

### Changes in monocyte phenotype within whole blood cultures

To establish the phenotype of circulating and cultured monocytes in CF and controls, we labelled fresh and 24 h cultured whole blood samples untreated or treated with M-CSF, GM-CSF, or GM-CSF+IL-4 for CD45 to identify haematopoietic cells and CD14 to identify monocytes, alongside CD11b, HLA-DR, CD206, and CD209 (see gating strategy in supplementary Figure 1).

**Figure 1.**
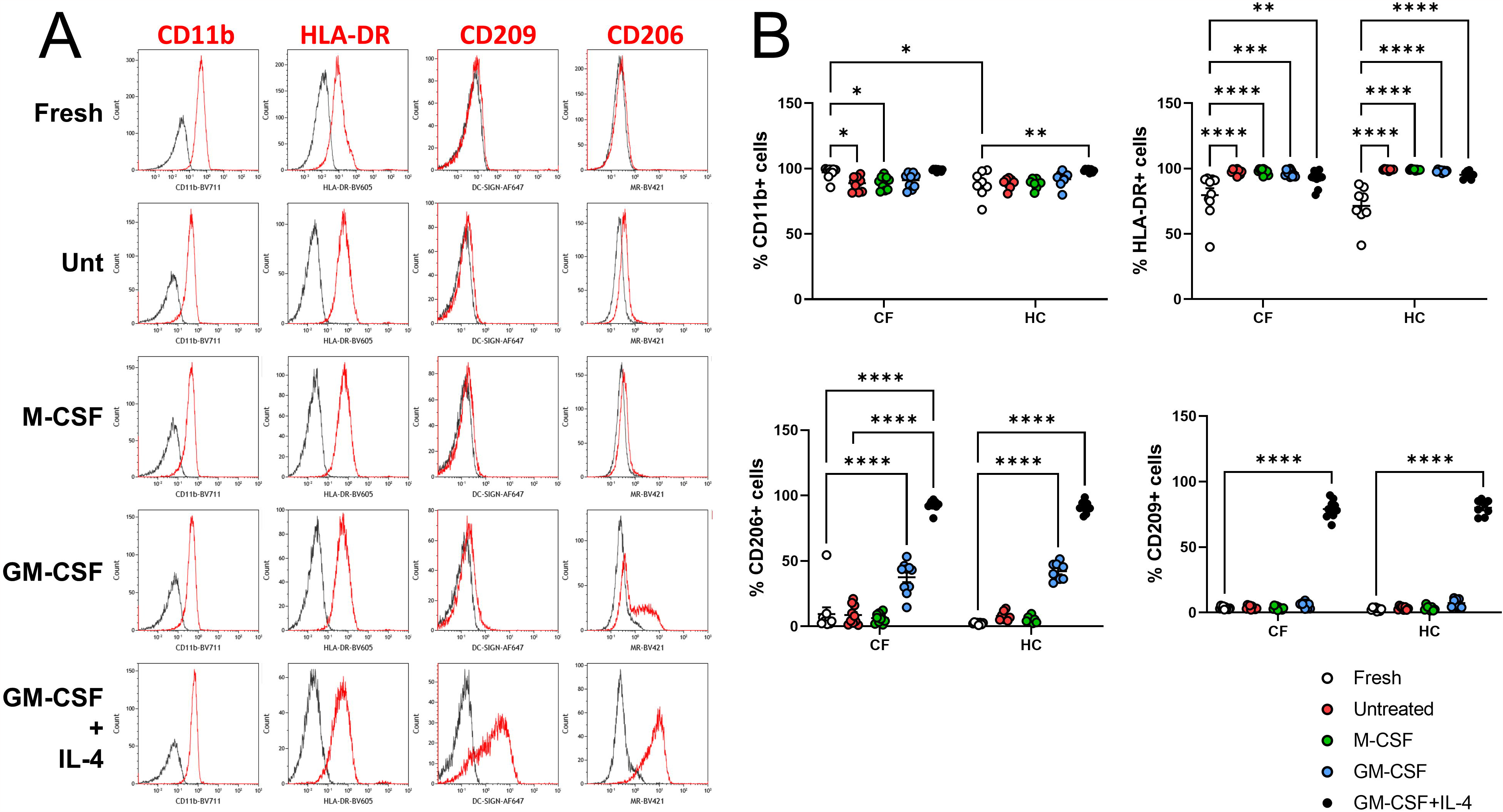
Analysis of percentages of CD45+CD14+ cells expressing CD11b, HLA-DR, CD206, and CD209 in CF and control fresh and cultured blood samples. Whole peripheral blood from CF and healthy control (HC) donors was either directly labelled and processed for flow cytometry or diluted 1:1 with complete RPMI medium and incubated for 24 h in the presence of M-CSF, GM-CSF, GM-CSF+IL-4, or no cytokines (Untreated, Unt), and then labelled and processed for flow cytometry. Both sets of samples were stained for CD45, CD14, CD11b, HLA-DR, CD206, and CD209. **A**. Representative histograms showing the expression of CD11b, HLA-DR, CD206, and CD209 on monocytes in fresh and cultured blood. Red lines represent the marker, while grey lines represent the corresponding isotype control. **B**. Monocytes from both donor groups consistently expressed CD11b and HLA-DR under all conditions with a higher percentage of CD11b+ cells observed in CF fresh blood compared to HC. Percentage of HLA-DR+ cells consistently significantly increased upon culture. The percentage of CD206+ cells was significantly increased in the presence of GM-CSF and GM-CSF+IL-4, while the percentage of CD209+ cells increased only in the presence of GM-CSF and IL-4. Graphs show mean ± standard error of mean (SEM) for n=10 CF donors and n=8 control donors. Data analysed using Two-Way ANOVA and Tukey’s multiple comparison test. (*) p<0.05; (**) p<0.01; (***) p< 0.0001; (****) p<0.0001.

CD45+CD14+ cells expressed CD11b and HLA-DR under all conditions (Figure 1). Mean percentage of CD11b+ cells in fresh blood samples was 97% for CF and 87% for HC, with this variation reaching statistical significance and indicating differences in circulating monocytes between CF and control individuals. There were minor changes in the percentages of cells expressing CD11b in CF and HC samples upon culture but no differences between both groups were detected. Percentage of HLA-DR+ cells was high under all conditions (>70% for CF and controls) with a significant upregulation and reduced variance upon culture, regardless of the presence of cytokines. CD206 was detected in very low percentage of cells in fresh, and untreated and M-CSF-treated cultures (6.32-9.42% for CF and 2.44-7.87% for HC) but was detected in approximately 40% of cells in the presence of GM-CSF (37.58% for CF and 42.16% for HC) and 90% in the presence of GM-CSF+IL-4 (92.68% for CF and 90.96% for HC). CD209 was detected in 80% of CD45+CD14+ cells in GM-CSF+IL-4 cultures (79.12 % for CF and 80.1% for HC) with very low percentages of CD209+ cells detected in fresh, and untreated, M-CSF-and GM-CSF-treated cultures (3.54-5.90% for CF and 2.64-7.32% for HC). We did not observe any differences in the percentages of cells expressing HLA-DR, CD206, or CD209 between CF and HC samples under any conditions.

### CD206 upregulation differs in CF monocytes and correlates with lung function

Analysis of rMFI for CD11b, HLA-DR, CD206, and CD209 in CD45+CD14+ cells (Figure 2) revealed that CD11b rMFI was comparable in fresh and cultured blood, although there was a trend towards increased expression in CF compared with HC samples under all conditions. HLA-DR rMFI values increased in response to GM-CSF+IL-4 and we observed no differences between CF and HC samples. CD206 rMFI was significantly upregulated by GM-CSF+IL-4 in CD45+CD14+ cells in CF and control cultures but only significantly upregulated by GM-CSF in control cultures. Further analysis of this finding using multiple unpaired T tests or Mann-Whitney tests correcting for multiple comparisons (Holm-Sidak method), showed that differences in GM-CSF samples between CF and controls were statistically significant [adjusted p value 0.0136 (T test) and 0.0068 (Mann-Whitney test)]. CD209 was significantly upregulated by GM-CSF+IL-4 in CF samples but not in controls. This is possibly due to the three high responders in the CF cohort.

**Figure 2.**
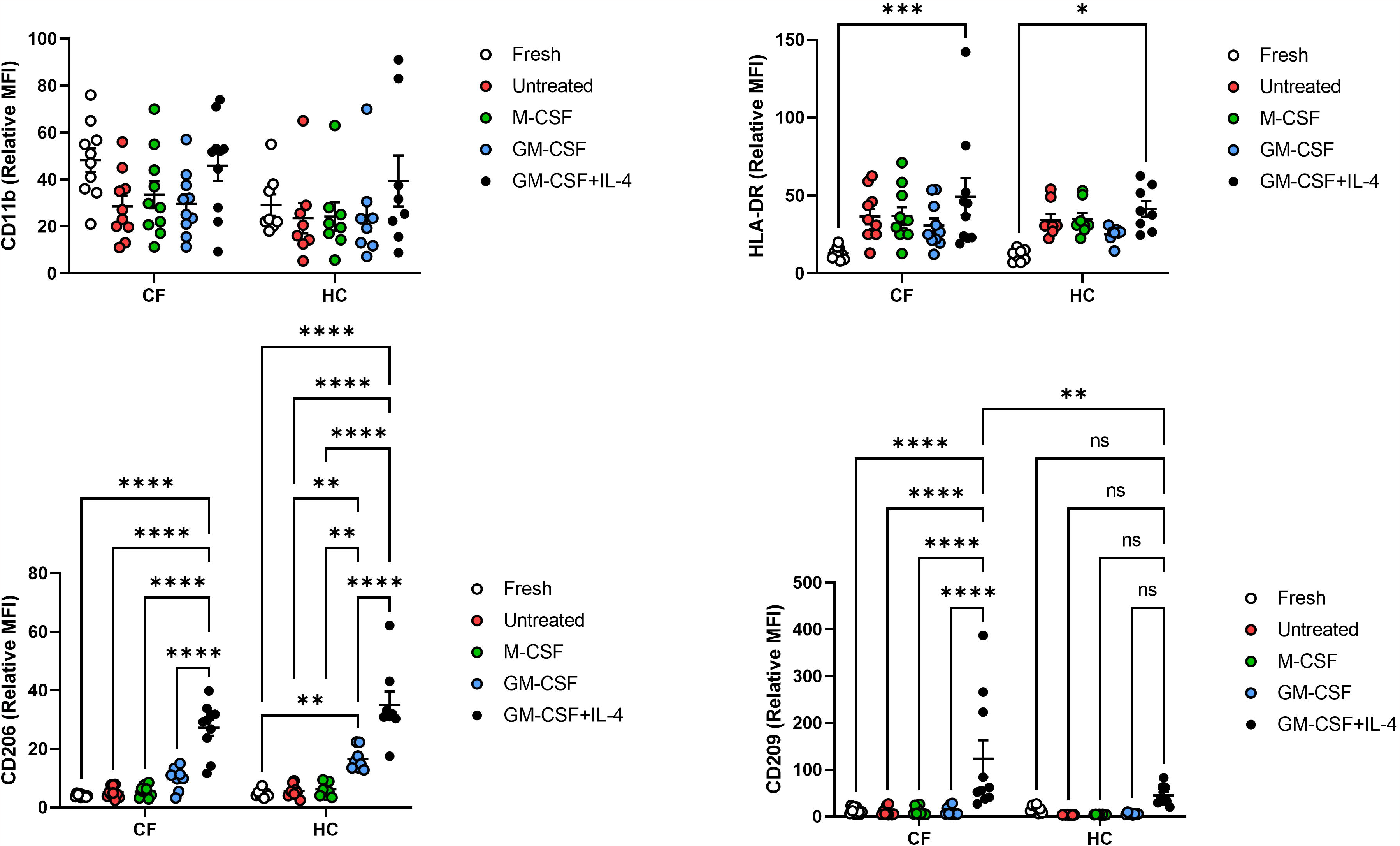
Analysis of CD11b, HLA-DR, CD206, and CD209 rMFI in CD45+CD14+ cells in CF and control fresh and cultured blood samples. Whole peripheral blood from CF and healthy control (HC) donors was either directly labelled and processed for flow cytometry or diluted 1:1 with complete RPMI medium and incubated for 24 h in the presence of M-CSF, GM-CSF, GM-CSF+IL-4, or no cytokines (Untreated, Unt), and then labelled and processed for flow cytometry. Both sets of samples were stained for CD45, CD14, CD11b, HLA-DR, CD206, and CD209. CD11b rMFI values were highly heterogeneous and no clear trend was observed regarding sample origin or treatment. HLA-DR rMFI displayed a clear trend towards upregulation upon culture, with this trend becoming statistically significant in GM-CSF+IL-4 samples. CD206 rMFI was significantly upregulated in CF and HC cultures in the presence of GM-CSF+IL-4. Upregulation in response to GM-CSF only reached significance in HC cultures. CD209 rMFI was only increased in GM-CSF+IL-4 cultures with this upregulation only reaching significance for CF samples. Data analysed using Two-Way ANOVA and Tukey’s multiple comparison test. (*) p<0.05; (**) p<0.01; (***) p< 0.0001; (****) p<0.0001.

To establish the potential clinical relevance of these findings we explored correlations between marker expression (as percentages of expressing cells and rMFI) and lung function. We observed a positive correlation between CD206 rMFI in CD45+CD14+ cells within whole blood cultures exposed to M-CSF, GM-CSF, or GM-CSF+IL-4 with lung function (FEV_1_) in CF donors (Figure 3). We found no such correlations for CD11b, HLA-DR, or CD209 (data not shown).

**Figure 3.**
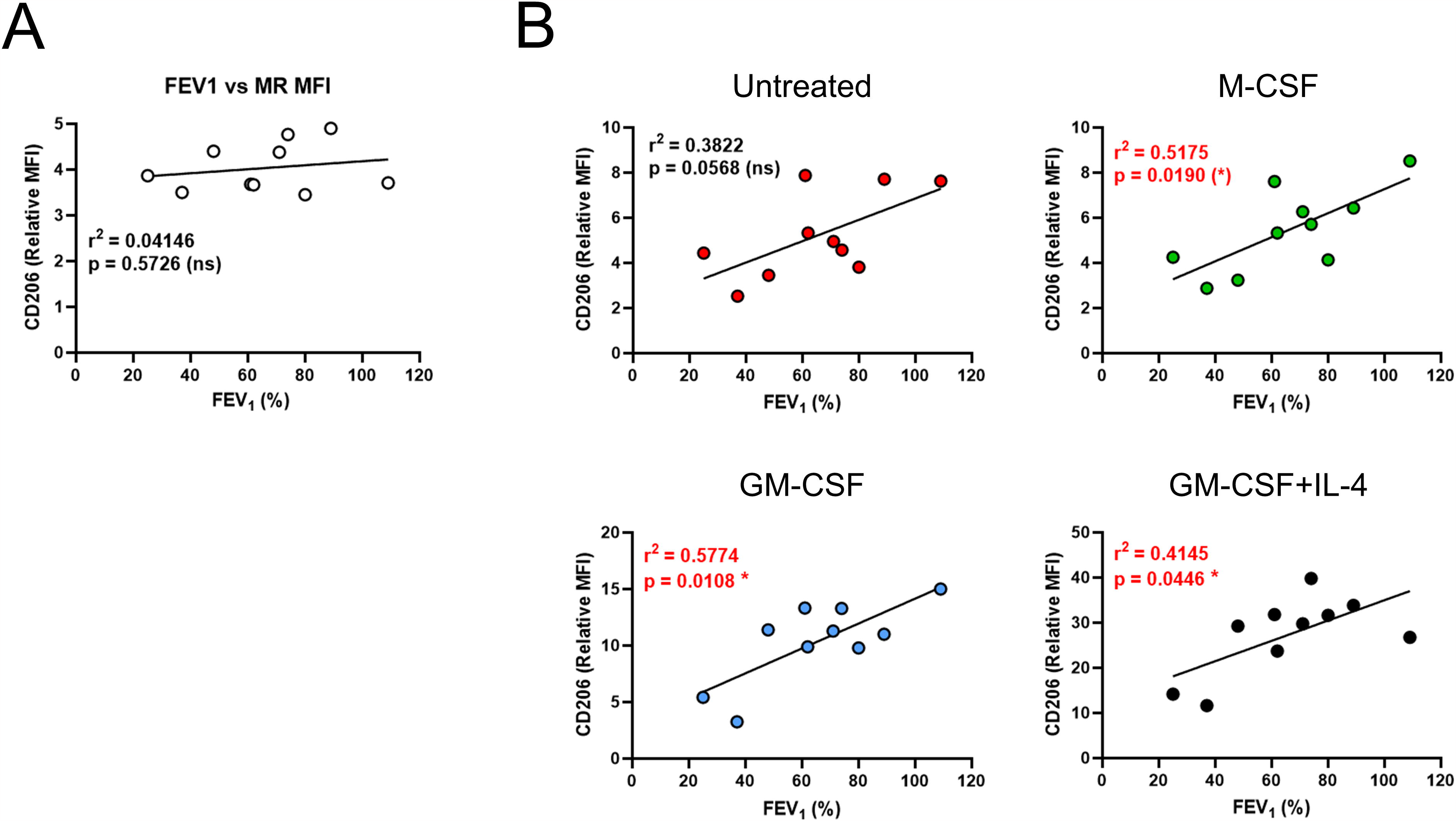
CD206 rMFI in CD45+CD14+ cells within 24 h whole blood cultures positively correlate with lung function in CF. Whole peripheral blood from CF donors was either directly labelled and processed for flow cytometry or diluted 1:1 with complete RPMI medium and incubated for 24 h in the presence of M-CSF, GM-CSF, GM-CSF+IL-4, or no cytokines (Untreated), and then labelled and processed for flow cytometry. All samples were stained for CD45, CD14, CD11b, HLA-DR, CD206 and CD209. N=10. Graphs show Pearson correlation analysis between lung function (FEV_1_) of CF donors and CD206 rMFI in CD45+CD14+ cells in fresh blood **(A)** and after 24 h culture **(B)**. Note differences in the scale of the Y axis for each condition.

## 4. Discussion

The aim of this proof of principle study was to establish the possibility of detecting differences in the phenotype of differentiating monocytes within the host environment between people with CF and controls. Within this experimental set up (whole blood cultures), monocytes are influenced by the mutations in CFTR (ΔF508 in this instance), alongside environmental factors including other circulating cells that can also respond to GM-CSF and/or IL-4, and circulating microbial products including endotoxin and quorum sensing molecules [18-21]. We stimulated the cultures with factors that promote homeostatic (M-CSF) and inflammatory and mucosal (GM-CSF) monocyte differentiation. There is extensive literature of how both cytokines affect monocytes and macrophages including upregulation of CD206 by GM-CSF in purified monocytes [22] and PBMC cultures [23]. Addition of GM-CSF was not an attempt to mimic inflammatory conditions in CF, indeed circulating levels of GM-CSF are low in CF [24, 25]. The aim was to compare how monocytes would behave in M-CSF-dominated vs. GM-CSF-dominated environments.

Circulating CF and control monocytes were CD206-, CD209-, CD11b+, and HLA-DR+. We noted a significant trend for higher percentage of CD11b+ monocytes in fresh blood samples from CF compared to control donors. CD11b forms the α chain of the integrin CD11b/CD18, which plays a critical role in leukocyte adhesion and transmigration amongst other functions [26]. Elevated CD11b levels in CF may be caused by the inflammatory environment [27]. HLA-DR was upregulated upon culture, particularly in presence of GM-CSF+IL-4, a combination widely used to generate monocyte-derived dendritic cells with high T cell activating capacity [15], but no differences were observed in HLA-DR expression between CF and HC samples.

CF has been characterised as a Th2/Th17 -dominated inflammatory disease [2]. Results from untreated and M-CSF-treated whole blood cultures indicate that the environment did not provide enough signals for induction of CD206 and CD209 expression after 24 h culture. CD206 and CD209 are both upregulated by IL-4 in human macrophages [28, 29]. The GM-CSF+IL-4 condition indicates that CF and HC monocytes were capable of upregulating CD206 and CD209 when suitably activated.

CD206 was upregulated only in 40% of monocytes by GM-CSF in 24 h cultures (both CF and HC). It is possible that this percentage would increase if cultures were maintained for longer. Intriguingly, in HC cultures, GM-CSF induced a significant increase in CD206 rMFI in monocytes compared to fresh, and untreated and M-CSF-treated cultures. This was not the case in CF despite both CF and HC cultures supporting similar CD206 rMFI upregulation in response to GM-CSF+IL-4. We speculate that CF monocytes might be less responsive to GM-CSF. Since GM-CSF is likely found at higher concentrations in the lung, recruited lung monocytes could display less CD206 in CF. This would agree with the reduced CD206 expression observed in a population of small CD14^+^, HLA-DR^+^, CD68^weak^ sputum macrophages expanded in CF compared to controls, which could correspond to recruited inflammatory monocytes [30]. These cells would represent a different population to alveolar macrophages shown to express high levels of CD206 [31]. Lung macrophages are of monocytic origin and upregulate CD206 upon differentiation [32]. Murphy *et al*. [33] observed a negative correlation between percentage of CD206+ cells within CD11b+ cells (used as marker of infiltrating monocytes) in sputum and bronchoalveolar lavage of CF patients regardless of *P. aeruginosa* infection status, with *P. aeruginosa* infection being associated with increased percentage of CD206+ cells. Our results showing significant positive correlations between lung function and CD206 rMFI in M-CSF, GM-CSF, and GM-CSF+IL-4-treated monocytes in CF complement these findings. Both support the notion that physiological conditions in CF influence CD206 expression.

Understanding the relationship between CD206 and clinical status in CF may provide new biomarkers and/or therapeutic interventions, and as such, deserves further investigation.

Lower CD206 upregulation in response to GM-CSF in CF monocytes could be caused by (i) reduced transcription because of substantial levels of endotoxin in CF plasma [20] or reduced cellular responses to GM-CSF in CF, or (ii) increased release of soluble CD206 (sCD206) which is enhanced in response different stimuli [34]. Reduced responsiveness to GM-CSF in CF might have implications for wound healing at mucosal sites [14]. CD206 can be released from cells as soluble form: sCD206 [35, 36]. Raised levels of sCD206 are found in inflammatory conditions [37], and sCD206 is of prognostic value in pulmonary tuberculosis [38] and community-acquired pneumonia [39]. Analysis of sCD206 in CF would clarify this point.

This pilot study has clear limitations such as the reduced number of samples and markers analysed, lack of longitudinal samples, and no quantification of serum biomarkers like levels of LPS or cytokines. Nevertheless, our results clearly pave the way for the exploitation of whole blood cultures to analyse monocyte differentiation in CF and other inflammatory diseases and assess effect of therapeutics such as CFTR-modulators on immune cells. These studies will benefit from increasing the range of parameters analysed informed by high-dimensional phenotyping, [3, 40, 41] that could be linked to serum biomarkers and sputum analysis.

## Data Availability

All data produced in the present work are contained in the manuscript

## Acknowledgements

This study was funded by UKRI-MRC grant [Award number MR/P001033/1] to Luisa Martinez-Pomares, Miguel Camara and Paul Williams, and the National Biofilms Innovation Centre (NBIC) which is an Innovation and Knowledge Centre funded by the Biotechnology and Biological Sciences Research Council, Innovate UK and Hartree Centre [Awards Numbers BB/R012415/1 to MC, LMP and PW and BB/X002950/1 to MC].

## References

1. Marteyn BS, Burgel PR, Meijer L, Witko-Sarsat V. Harnessing Neutrophil Survival Mechanisms during Chronic Infection by Pseudomonas aeruginosa: Novel Therapeutic Targets to Dampen Inflammation in Cystic Fibrosis. Front Cell Infect Microbiol. 2017;7:243. Epub 2017/07/18. doi: 10.3389/fcimb.2017.00243. PubMed PMID: 28713772; PubMed Central PMCID: PMCPMC5492487.

2. Tiringer K, Treis A, Fucik P, Gona M, Gruber S, Renner S, et al. A Th17- and Th2-skewed cytokine profile in cystic fibrosis lungs represents a potential risk factor for Pseudomonas aeruginosa infection. Am J Respir Crit Care Med. 2013;187(6):621–9. Epub 2013/01/12. doi: 10.1164/rccm.201206-1150OC. PubMed PMID: 23306544.

3. Horsley AR, Davies JC, Gray RD, Macleod KA, Donovan J, Aziz ZA, et al. Changes in physiological, functional and structural markers of cystic fibrosis lung disease with treatment of a pulmonary exacerbation. Thorax. 2013;68(6):532–9. Epub 2013/02/12. doi: 10.1136/thoraxjnl-2012-202538. PubMed PMID: 23396354.

4. Singh S, Barr H, Liu YC, Robins A, Heeb S, Williams P, et al. Granulocyte-macrophage colony stimulatory factor enhances the pro-inflammatory response of interferon-gamma-treated macrophages to Pseudomonas aeruginosa infection. PLoS One. 2015;10(2):e0117447. Epub 2015/02/24. doi: 10.1371/journal.pone.0117447. PubMed PMID: 25706389; PubMed Central PMCID: PMCPMC4338139.

5. Tarique AA, Sly PD, Holt PG, Bosco A, Ware RS, Logan J, et al. CFTR-dependent defect in alternatively-activated macrophages in cystic fibrosis. J Cyst Fibros. 2017;16(4):475–82. Epub 2017/04/22. doi: 10.1016/j.jcf.2017.03.011. PubMed PMID: 28428011.

6. Cavinato L, Luly FR, Pastore V, Chiappetta D, Sangiorgi G, Ferrara E, et al. Elexacaftor/tezacaftor/ivacaftor corrects monocyte microbicidal deficiency in cystic fibrosis. Eur Respir J. 2023;61(4). Epub 2022/12/02. doi: 10.1183/13993003.00725-2022. PubMed PMID: 36455959; PubMed Central PMCID: PMCPMC10066567.

7. Gabillard-Lefort C, Casey M, Glasgow AMA, Boland F, Kerr O, Marron E, et al. Trikafta Rescues CFTR and Lowers Monocyte P2X7R-induced Inflammasome Activation in Cystic Fibrosis. Am J Respir Crit Care Med. 2022;205(7):783–94. Epub 2022/01/13. doi: 10.1164/rccm.202106-1426OC. PubMed PMID: 35021019.

8. Oz HH, Cheng EC, Di Pietro C, Tebaldi T, Biancon G, Zeiss C, et al. Recruited monocytes/macrophages drive pulmonary neutrophilic inflammation and irreversible lung tissue remodeling in cystic fibrosis. Cell Rep. 2022;41(11):111797. Epub 2022/12/15. doi: 10.1016/j.celrep.2022.111797. PubMed PMID: 36516754; PubMed Central PMCID: PMCPMC9833830.

9. Damsgaard CT, Lauritzen L, Calder PC, Kjaer TM, Frokiaer H. Whole-blood culture is a valid low-cost method to measure monocytic cytokines - a comparison of cytokine production in cultures of human whole-blood, mononuclear cells and monocytes. J Immunol Methods. 2009;340(2):95–101. Epub 2008/11/13. doi: 10.1016/j.jim.2008.10.005. PubMed PMID: 19000693.

10. Duffy D, Rouilly V, Libri V, Hasan M, Beitz B, David M, et al. Functional analysis via standardized whole-blood stimulation systems defines the boundaries of a healthy immune response to complex stimuli. Immunity. 2014;40(3):436–50. Epub 2014/03/25. doi: 10.1016/j.immuni.2014.03.002. PubMed PMID: 24656047.

11. Ait-Belkacem I, Cartagena Garcia C, Millet-Wallisky E, Izquierdo N, Loosveld M, Arnoux I, et al. SARS-CoV-2 spike protein induces a differential monocyte activation that may contribute to age bias in COVID-19 severity. Sci Rep. 2022;12(1):20824. Epub 2022/12/03. doi: 10.1038/s41598-022-25259-2. PubMed PMID: 36460710; PubMed Central PMCID: PMCPMC9716544 employees. All other authors declare no competing interests.

12. Hamilton JA. Colony-stimulating factors in inflammation and autoimmunity. Nat Rev Immunol. 2008;8(7):533–44. Epub 2008/06/14. doi: 10.1038/nri2356. PubMed PMID: 18551128.

13. Gschwend J, Sherman SPM, Ridder F, Feng X, Liang HE, Locksley RM, et al. Alveolar macrophages rely on GM-CSF from alveolar epithelial type 2 cells before and after birth. J Exp Med. 2021;218(10). Epub 2021/08/26. doi: 10.1084/jem.20210745. PubMed PMID: 34431978; PubMed Central PMCID: PMCPMC8404471.

14. Dabritz J, Weinhage T, Varga G, Wirth T, Walscheid K, Brockhausen A, et al. Reprogramming of monocytes by GM-CSF contributes to regulatory immune functions during intestinal inflammation. J Immunol. 2015;194(5):2424–38. Epub 2015/02/06. doi: 10.4049/jimmunol.1401482. PubMed PMID: 25653427.

15. Sallusto F, Lanzavecchia A. Efficient presentation of soluble antigen by cultured human dendritic cells is maintained by granulocyte/macrophage colony-stimulating factor plus interleukin 4 and downregulated by tumor necrosis factor alpha. J Exp Med. 1994;179(4):1109–18. Epub 1994/04/01. doi: 10.1084/jem.179.4.1109. PubMed PMID: 8145033; PubMed Central PMCID: PMCPMC2191432.

16. Singh S, Almuhanna Y, Alshahrani MY, Lowman DW, Rice PJ, Gell C, et al. Carbohydrates from Pseudomonas aeruginosa biofilms interact with immune C-type lectins and interfere with their receptor function. NPJ Biofilms Microbiomes. 2021;7(1):87. Epub 2021/12/10. doi: 10.1038/s41522-021-00257-w. PubMed PMID: 34880222; PubMed Central PMCID: PMCPMC8655052.

17. Kapellos TS, Bonaguro L, Gemund I, Reusch N, Saglam A, Hinkley ER, et al. Human Monocyte Subsets and Phenotypes in Major Chronic Inflammatory Diseases. Front Immunol. 2019;10:2035. Epub 2019/09/24. doi: 10.3389/fimmu.2019.02035. PubMed PMID: 31543877; PubMed Central PMCID: PMCPMC6728754.

18. Barr HL, Halliday N, Barrett DA, Williams P, Forrester DL, Peckham D, et al. Diagnostic and prognostic significance of systemic alkyl quinolones for P. aeruginosa in cystic fibrosis: A longitudinal study. J Cyst Fibros. 2017;16(2):230–8. Epub 2016/10/25. doi: 10.1016/j.jcf.2016.10.005. PubMed PMID: 27773591; PubMed Central PMCID: PMCPMC5345566.

19. Barr HL, Halliday N, Camara M, Barrett DA, Williams P, Forrester DL, et al. Pseudomonas aeruginosa quorum sensing molecules correlate with clinical status in cystic fibrosis. Eur Respir J. 2015;46(4):1046–54. Epub 2015/05/30. doi: 10.1183/09031936.00225214. PubMed PMID: 26022946; PubMed Central PMCID: PMCPMC4589431 this article at erj.ersjournals.com.

20. del Campo R, Martinez E, del Fresno C, Alenda R, Gomez-Pina V, Fernandez-Ruiz I, et al. Translocated LPS might cause endotoxin tolerance in circulating monocytes of cystic fibrosis patients. PLoS One. 2011;6(12):e29577. Epub 2012/01/05. doi: 10.1371/journal.pone.0029577. PubMed PMID: 22216320; PubMed Central PMCID: PMCPMC3247277.

21. Zain NMM, Webb K, Stewart I, Halliday N, Barrett DA, Nash EF, et al. 2-Alkyl-4-quinolone quorum sensing molecules are biomarkers for culture-independent Pseudomonas aeruginosa burden in adults with cystic fibrosis. J Med Microbiol. 2021;70(10). Epub 2021/10/02. doi: 10.1099/jmm.0.001420. PubMed PMID: 34596013; PubMed Central PMCID: PMCPMC8604169.

22. Rodriguez RM, Suarez-Alvarez B, Lavin JL, Ascension AM, Gonzalez M, Lozano JJ, et al. Signal Integration and Transcriptional Regulation of the Inflammatory Response Mediated by the GM-/M-CSF Signaling Axis in Human Monocytes. Cell Rep. 2019;29(4):860–72 e5. Epub 2019/10/24. doi: 10.1016/j.celrep.2019.09.035. PubMed PMID: 31644909.

23. Tan-Garcia A, Lai F, Sheng Yeong JP, Irac SE, Ng PY, Msallam R, et al. Liver fibrosis and CD206(+) macrophage accumulation are suppressed by anti-GM-CSF therapy. JHEP Rep. 2020;2(1):100062. Epub 2020/02/11. doi: 10.1016/j.jhepr.2019.11.006. PubMed PMID: 32039403; PubMed Central PMCID: PMCPMC7005658.

24. Castellani S, D’Oria S, Diana A, Polizzi AM, Di Gioia S, Mariggio MA, et al. G-CSF and GM-CSF Modify Neutrophil Functions at Concentrations found in Cystic Fibrosis. Sci Rep. 2019;9(1):12937. Epub 2019/09/12. doi: 10.1038/s41598-019-49419-z. PubMed PMID: 31506515; PubMed Central PMCID: PMCPMC6736848.

25. Moser C, Jensen PO, Pressler T, Frederiksen B, Lanng S, Kharazmi A, et al. Serum concentrations of GM-CSF and G-CSF correlate with the Th1/Th2 cytokine response in cystic fibrosis patients with chronic Pseudomonas aeruginosa lung infection. APMIS. 2005;113(6):400–9. Epub 2005/07/06. doi: 10.1111/j.1600-0463.2005.apm_142.x. PubMed PMID: 15996157.

26. Villanueva V, Li X, Jimenez V, Faridi HM, Gupta V. CD11b agonists offer a novel approach for treating lupus nephritis. Transl Res. 2022;245:41–54. Epub 2022/03/16. doi: 10.1016/j.trsl.2022.03.001. PubMed PMID: 35288363; PubMed Central PMCID: PMCPMC9167730.

27. Dadfar E, Jacobson SH, Lundahl J. Newly recruited human monocytes have a preserved responsiveness towards bacterial peptides in terms of CD11b up-regulation and intracellular hydrogen peroxide production. Clin Exp Immunol. 2007;148(3):573–82. Epub 2007/03/28. doi: 10.1111/j.1365-2249.2007.03373.x. PubMed PMID: 17386075; PubMed Central PMCID: PMCPMC1941923.

28. Jayme TS, Leung G, Wang A, Workentine ML, Rajeev S, Shute A, et al. Human interleukin-4-treated regulatory macrophages promote epithelial wound healing and reduce colitis in a mouse model. Sci Adv. 2020;6(23):eaba4376. Epub 2020/06/18. doi: 10.1126/sciadv.aba4376. PubMed PMID: 32548267; PubMed Central PMCID: PMCPMC7274799.

29. Lugo-Villarino G, Troegeler A, Balboa L, Lastrucci C, Duval C, Mercier I, et al. The C-Type Lectin Receptor DC-SIGN Has an Anti-Inflammatory Role in Human M(IL-4) Macrophages in Response to Mycobacterium tuberculosis. Front Immunol. 2018;9:1123. Epub 2018/06/28. doi: 10.3389/fimmu.2018.01123. PubMed PMID: 29946317; PubMed Central PMCID: PMCPMC6006465.

30. Wright AK, Rao S, Range S, Eder C, Hofer TP, Frankenberger M, et al. Pivotal Advance: Expansion of small sputum macrophages in CF: failure to express MARCO and mannose receptors. J Leukoc Biol. 2009;86(3):479–89. Epub 2009/05/01. doi: 10.1189/jlb.1108699. PubMed PMID: 19403625.

31. Desch AN, Gibbings SL, Goyal R, Kolde R, Bednarek J, Bruno T, et al. Flow Cytometric Analysis of Mononuclear Phagocytes in Nondiseased Human Lung and Lung-Draining Lymph Nodes. Am J Respir Crit Care Med. 2016;193(6):614–26. Epub 2015/11/10. doi: 10.1164/rccm.201507-1376OC. PubMed PMID: 26551758; PubMed Central PMCID: PMCPMC4824940.

32. Evren E, Ringqvist E, Tripathi KP, Sleiers N, Rives IC, Alisjahbana A, et al. Distinct developmental pathways from blood monocytes generate human lung macrophage diversity. Immunity. 2021;54(2):259–75 e7. Epub 2021/01/01. doi: 10.1016/j.immuni.2020.12.003. PubMed PMID: 33382972.

33. Murphy BS, Bush HM, Sundareshan V, Davis C, Hagadone J, Cory TJ, et al. Characterization of macrophage activation states in patients with cystic fibrosis. J Cyst Fibros. 2010;9(5):314–22. Epub 2010/06/24. doi: 10.1016/j.jcf.2010.04.006. PubMed PMID: 20570573.

34. Nielsen MC, Andersen MN, Rittig N, Rodgaard-Hansen S, Gronbaek H, Moestrup SK, et al. The macrophage-related biomarkers sCD163 and sCD206 are released by different shedding mechanisms. J Leukoc Biol. 2019;106(5):1129–38. Epub 2019/06/27. doi: 10.1002/JLB.3A1218-500R. PubMed PMID: 31242338.

35. Jordens R, Thompson A, Amons R, Koning F. Human dendritic cells shed a functional, soluble form of the mannose receptor. Int Immunol. 1999;11(11):1775–80. Epub 1999/11/05. doi: 10.1093/intimm/11.11.1775. PubMed PMID: 10545481.

36. Martinez-Pomares L, Mahoney JA, Kaposzta R, Linehan SA, Stahl PD, Gordon S. A functional soluble form of the murine mannose receptor is produced by macrophages in vitro and is present in mouse serum. J Biol Chem. 1998;273(36):23376–80. Epub 1998/08/29. doi: 10.1074/jbc.273.36.23376. PubMed PMID: 9722572.

37. De Vlieger G, Vanhorebeek I, Wouters PJ, Derese I, Casaer MP, Debaveye Y, et al. The soluble mannose receptor (sMR/sCD206) in critically ill patients with invasive fungal infections, bacterial infections or non-infectious inflammation: a secondary analysis of the EPaNIC RCT. Crit Care. 2019;23(1):270. Epub 2019/08/04. doi: 10.1186/s13054-019-2549-8. PubMed PMID: 31375142;PubMed Central PMCID: PMCPMC6679534.

38. Suzuki Y, Shirai M, Asada K, Yasui H, Karayama M, Hozumi H, et al. Macrophage mannose receptor, CD206, predict prognosis in patients with pulmonary tuberculosis. Sci Rep. 2018;8(1):13129. Epub 2018/09/05. doi: 10.1038/s41598-018-31565-5. PubMed PMID: 30177769; PubMed Central PMCID: PMCPMC6120933.

39. Tsuchiya K, Suzuki Y, Yoshimura K, Yasui H, Karayama M, Hozumi H, et al. Macrophage Mannose Receptor CD206 Predicts Prognosis in Community-acquired Pneumonia. Sci Rep. 2019;9(1):18750. Epub 2019/12/12. doi: 10.1038/s41598-019-55289-2. PubMed PMID: 31822747; PubMed Central PMCID: PMCPMC6904766.

40. Sander J, Schmidt SV, Cirovic B, McGovern N, Papantonopoulou O, Hardt AL, et al. Cellular Differentiation of Human Monocytes Is Regulated by Time-Dependent Interleukin-4 Signaling and the Transcriptional Regulator NCOR2. Immunity. 2017;47(6):1051–66 e12. Epub 2017/12/21. doi: 10.1016/j.immuni.2017.11.024. PubMed PMID: 29262348; PubMed Central PMCID: PMCPMC5772172.

41. Schupp JC, Khanal S, Gomez JL, Sauler M, Adams TS, Chupp GL, et al. Single-Cell Transcriptional Archetypes of Airway Inflammation in Cystic Fibrosis. Am J Respir Crit Care Med. 2020;202(10):1419–29. Epub 2020/07/01. doi: 10.1164/rccm.202004-0991OC. PubMed PMID: 32603604; PubMed Central PMCID: PMCPMC7667912.

