## Supplementary Information for "CD206 upregulation in monocytes within whole blood cultures correlates with lung function in Cystic Fibrosis: a pilot study"

**Supplementary Table 1. Reagents and consumables used in this study.**

| **Item** | **Manufacturer/Supplier** | **Catalogue number** |
| --- | --- | --- |
| BD Vacutainer® Citrate Tubes (PET/PP) 2.7mL | BD | 363095 |
| OptiLyse C | Beckman Coulter | A11895 |
| 16% Paraformaldehyde aqueous solution, EM grade | Electron Microscopy Sciences | 15710-S |
| Dulbecco′s Phosphate Buffered Saline (PBS) | Sigma-Aldrich | D8537 |
| RPMI-1640 Medium | Sigma-Aldrich | R0883 |
| Human AB Serum | Sigma-Aldrich | H4522 |
| GlutaMAX | Gibco | 35050-038 |
| HEPES | Gibco | 15630-056 |
| Ultra-low attachment 24-well plates | Costar | 3473 |
| Recombinant human M-CSF | R&D Systems or | 216-MCC-010/CF |
|  | Miltenyi Biotec | 130-096-492 |
| Recombinant human GM-CSF | Miltenyi Biotec | 130-093-864 |
| Recombinant human IL-4 | Miltenyi Biotec | 130-093-920 |

**Supplementary Table 2. Antibodies used in this study.** Catalogue number and manufacturer for each antibody is given in parentheses.

| **Marker** | **Antibody** | **Isotype control** |
| --- | --- | --- |
| CD45 | PerCP/Cy5.5 anti-human CD45 antibody *(368503, BioLegend)* | PerCP/Cy5.5 Mouse IgG1 κ isotype control antibody *(400149, BioLegend)* |
| CD14 | PE-Cy7 anti-human CD14 antibody, *(A22331, Beckman Coulter)* | PE-Cy7 Mouse IgG2 α isotype control antibody *(A12692, Beckman Coulter)* |
| HLA-DR | Brilliant Violet™ 605 anti-human HLA-DR antibody *(307639, BioLegend)* | Brilliant Violet™ 605 Mouse IgG2 α,κ isotype control antibody *(400269, BioLegend)* |
| CD11b | Brilliant Violet™ 711 anti-human CD11b antibody *(301344, BioLegend)* | Brilliant Violet™ 711 Mouse IgG1 κ isotype control antibody *(400168, BioLegend)* |
| MR  (CD206) | Brilliant Violet™ 421 anti-human CD206 antibody *(321125, BioLegend)* | Brilliant Violet™ 421 Mouse IgG1 κ isotype control antibody *(400158, BioLegend)* |
| DC-SIGN  (CD209) | Alexa Fluor® 647 anti-human CD209 antibody *(330112, BioLegend)* | Alexa Fluor® 647 Mouse IgG2 α,κ isotype control antibody *(400234, BioLegend)* |

**Supplementary Figure 1. Gating strategy employed to identify monocytes (CD45+CD14+ cells) in fresh and 24 h whole blood cultures.**

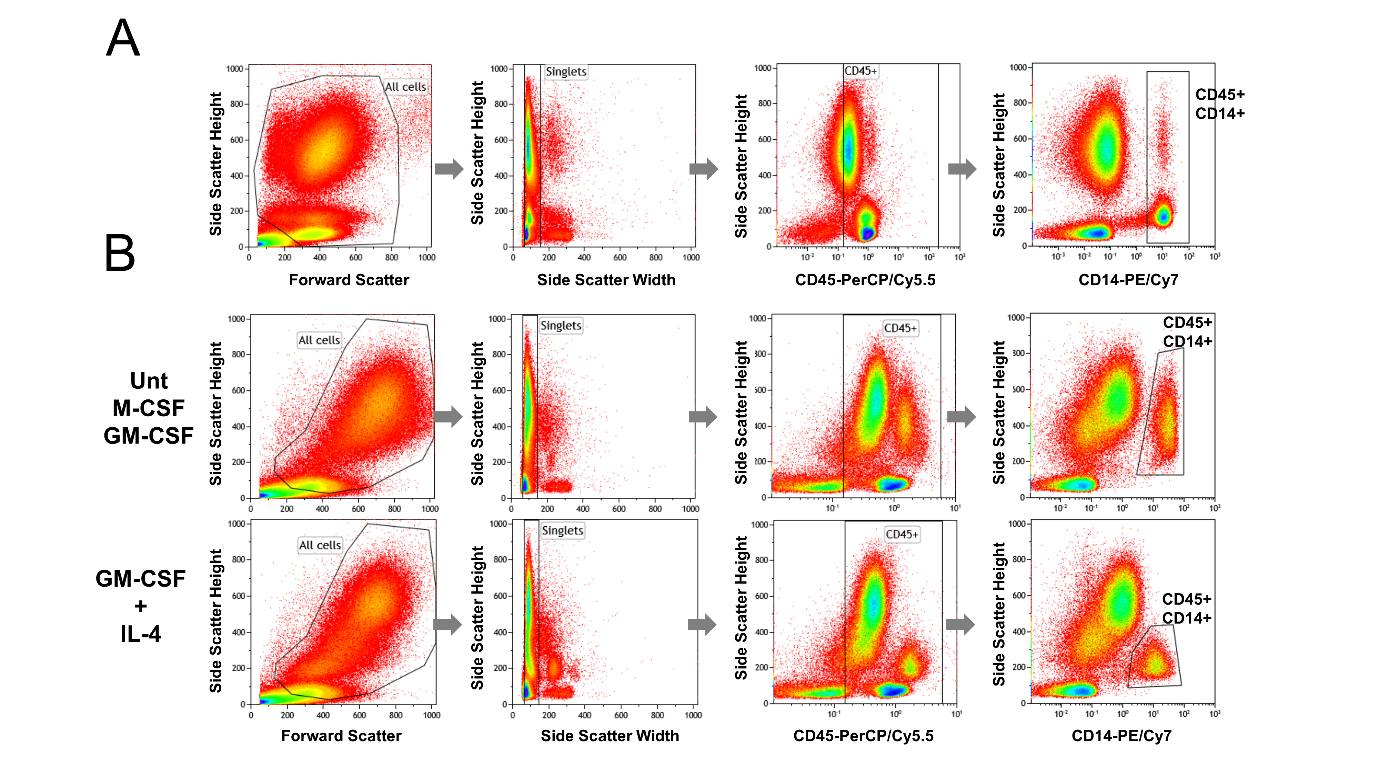

Whole peripheral blood from 10 CF and 8 control donors was stained for CD45, CD14, CD206, CD209, CD11b, and HLA-DR (A) or diluted 1:1 with complete RPMI medium and incubated for 24 h in the presence of M-CSF, GM-CSF, GM-CSF+IL-4, or no cytokines (Untreated, Unt). After incubation, whole blood cultures were stained for CD45, CD14, CD206, CD209, CD11b, and HLA-DR (B). CD45+ CD14+ cells within the Singlets gate were selected for analysis of CD11b, HLA-DR, CD206 and CD209 expression. CD45+CD14+ cells showed differences in the Side Scatter characteristics upon culture. In fresh blood, we noted two populations of CD14+ cells with different granularity. In untreated, M-CSF- and GM-CSF-treated cultures, the Side Scatter of CD45+CD14+ cells were comparable and showed a single heterogeneous population. In comparison, CD45+CD14+ cells in GM-CSF+IL-4-treated cultures displayed lower Side Scatter and were more homogeneous.
